# Cohort profile: OpenPROMPT

**DOI:** 10.1101/2023.12.21.23300369

**Authors:** Alasdair D Henderson, Oliver Carlile, Iain Dillingham, Ben FC Butler-Cole, Keith Tomlin, Mark Jit, Laurie A Tomlinson, Michael Marks, Andrew Briggs, Liang-Yu Lin, Chris Bates, John Parry, Sebastian CJ Bacon, Ben Goldacre, Amir Mehrkar, The OpenSAFELY Collaborative, Emily Herrett, Rosalind M Eggo

**Author notes:** Contributed equally (Emily Herrett, Rosalind M Eggo).

## Abstract

OpenPROMPT is a cohort of individuals with longitudinal patient reported questionnaire data and linked to routinely collected health data from primary and secondary care. Data were collected between November 2022 and October 2023 in England. OpenPROMPT was designed to measure the impact of long COVID on health-related quality-of-life (HRQoL). With the approval of NHS England we collected responses from 7,574 individuals, with detailed questionnaire responses from 6,337 individuals who responded using a smartphone app. Data were collected from each participant over 90 days at 30-day intervals using questionnaires to ask about HRQoL, productivity and symptoms of long COVID. Responses from the majority of OpenPROMPT (6,006; 79.3%) were linked to participants’ existing health records from primary care, secondary care, COVID-19 testing and vaccination data. Analysis takes place using the OpenSAFELY data analysis platform which provides a secure software interface allowing the analysis of pseudonymized primary care patient records from England. OpenPROMPT can currently be used to estimate the impact of long COVID on HRQoL, and because of the linkage within OpenSAFELY, the data from OpenPROMPT can be used to enrich routinely collected records in further research by approved researchers on behalf of NHS England.

**Lay summary:** OpenPROMPT is a study which used a phone app to conduct a longitudinal survey aimed at measuring the health related quality of life of people living with long COVID. The study recruited participants between November 2022 and July 2023 and followed them up for 90 days. The key advantage of this study is that the responses are linked to the individual’s personal health records, so we have access to much more data than the questionnaire responses alone.

Here, we summarised who has used the app, how much data has been collected and the quality of the data. We also provide details to document how and why the data were collected so that the data can be used by other researchers in the future. This will maximise the benefit of this study, and ensure that the time invested by participants is put to best use.

In this study we aimed to provide lots of important information about how many people are involved, how much information we have about them, their age, where they live, and how healthy they are. Finally, for certain variables we compared the responses people recorded in the app with what is kept on their electronic record to see if they agree or disagree.

**Key features:**

- OpenPROMPT is a cohort of individuals with longitudinal patient reported questionnaire data and linked to routinely collected health data from primary and secondary care.
- With the approval of NHS England we collected responses from 7,574 individuals, with detailed questionnaire responses from 6,337 individuals who responded using a smartphone app.
- Data were collected from each participant over 90 days at 30-day intervals using questionnaires to ask about HRQoL, productivity and symptoms of long COVID.
- Responses from the majority of OpenPROMPT (6,006; 79.3%) were linked to participants’ existing health records from primary care, secondary care, COVID-19 testing and vaccination data.
- OpenPROMPT can currently be used to estimate the impact of long COVID on HRQoL, and because of the linkage within OpenSAFELY, the data from OpenPROMPT can be used to enrich routinely collected records in further research by approved researchers on behalf of NHS England.

## Why was the cohort set up?

OpenPROMPT is a longitudinal prospective cohort of participant-recorded healthcare outcomes among UK adults, collected during the COVID-19 pandemic (November 2022 - July 2023). OpenPROMPT was funded by the National Institute for Healthcare research (NIHR) to better understand the impact of long COVID on health-related quality-of-life (HRQoL). Long COVID, here defined as symptoms consistent with SARS-COV-2 that persist for 12 weeks following an acute infection (1,2), has emerged as one of the most costly impacts of the pandemic in the UK (3) and globally (4,5). In the UK, the Office for National Statistics regularly collected information on the number and impact of persistent Covid-19 symptoms and as of November 2022 there were an estimated 2.1 million people with long COVID, 73% of whom reported that the symptoms adversely affected day-to-day activities (6).

Electronic health records (EHRs) are a valuable research tool to better understand the impact of health conditions. They have contributed to better understanding of COVID-19 fatalities (7,8), infection risk (9,10), the effectiveness of treatments (11,12) and vaccinations (13–15). However, long COVID in EHRs is poorly captured (16), and even in those that have a record this binary categorisation does not reflect the variety of experiences in those with long COVID, which may be better captured by patient-reported outcomes. Studies have previously used EHRs to study long COVID (16–19), and others have collected patient-reported outcome measurements (20–24), however OpenPROMPT is the first study of its kind to merge these two data sources in a trusted research environment (25).

The gold standard metric to quantify the impact of a disease is the quality adjusted life year (QALY), which takes into account impacts on morbidity as well as mortality. QALYs are used in National Health Service (NHS) resource planning to make healthcare decisions, evaluate and prioritise interventions, such as vaccination boosters for COVID-19. We therefore established OpenPROMPT to measure long COVID impacts using (1) standardised measures of changes in patient HRQoL (including physical and mental health effects) using the EuroQoL EQ-5D instrument (26) and (2) formal quantification of the healthcare burden of the condition and its treatment. We embedded patient and public involvement throughout OpenPROMPT to ensure that patient input on the effects of long COVID were included, and that possible concerns about data security were respected.

## Who is in the cohort

### Study design and setting

The protocol for data collection for OpenPROMPT was pre-specified and details have been previously published (27). Briefly, OpenPROMPT is a cohort study among adults in the UK, which recruited between November 2022 and July 2023. This study was conducted in collaboration with TPP, a general practitioner (GP) software provider. TPP’s software system is called SystmOne and holds patient electronic health records for approximately 40% of the population of England. Their in-house smartphone app, *Airmid* (28), has been developed as a patient-facing mobile application, and links directly to SystmOne records. It was created to allow appointment booking, for patients to access their own medical records, and for patients to input their own data to feed into their medical record and personal health record. We used Airmid’s Research Module to host bespoke questionnaires to assess quality of life in respondents. Any individual can download the Airmid app, even if their GP does not use TPP software, and access to Airmid and SystmOne was provided by TPP for no charge.

### Inclusion and exclusion criteria

Any adult with an NHS number was eligible to self-enrol in the study, irrespective of history of COVID-19 infection. There was an implicit exclusion criteria that participants needed to own a smartphone and be able to download and use the Airmid app. All responses to OpenPROMPT questionnaires are available for analysis, however access to linked primary care records is only available from those registered with a TPP practice.

### Linked healthcare records

OpenPROMPT responses are stored within the EHR vendor’s highly secure data centre and can be securely accessed through OpenSAFELY, a data analytics platform created by our team on behalf of NHS England to address urgent COVID-19 research questions (https://opensafely.org). If the respondent is registered with a TPP practice then their responses can be linked to other digital healthcare records within OpenSAFELY, including hospitalisations, prescriptions, GP appointments, diagnoses and COVID-19 test results (**Figure 1**).

**Figure 1:**
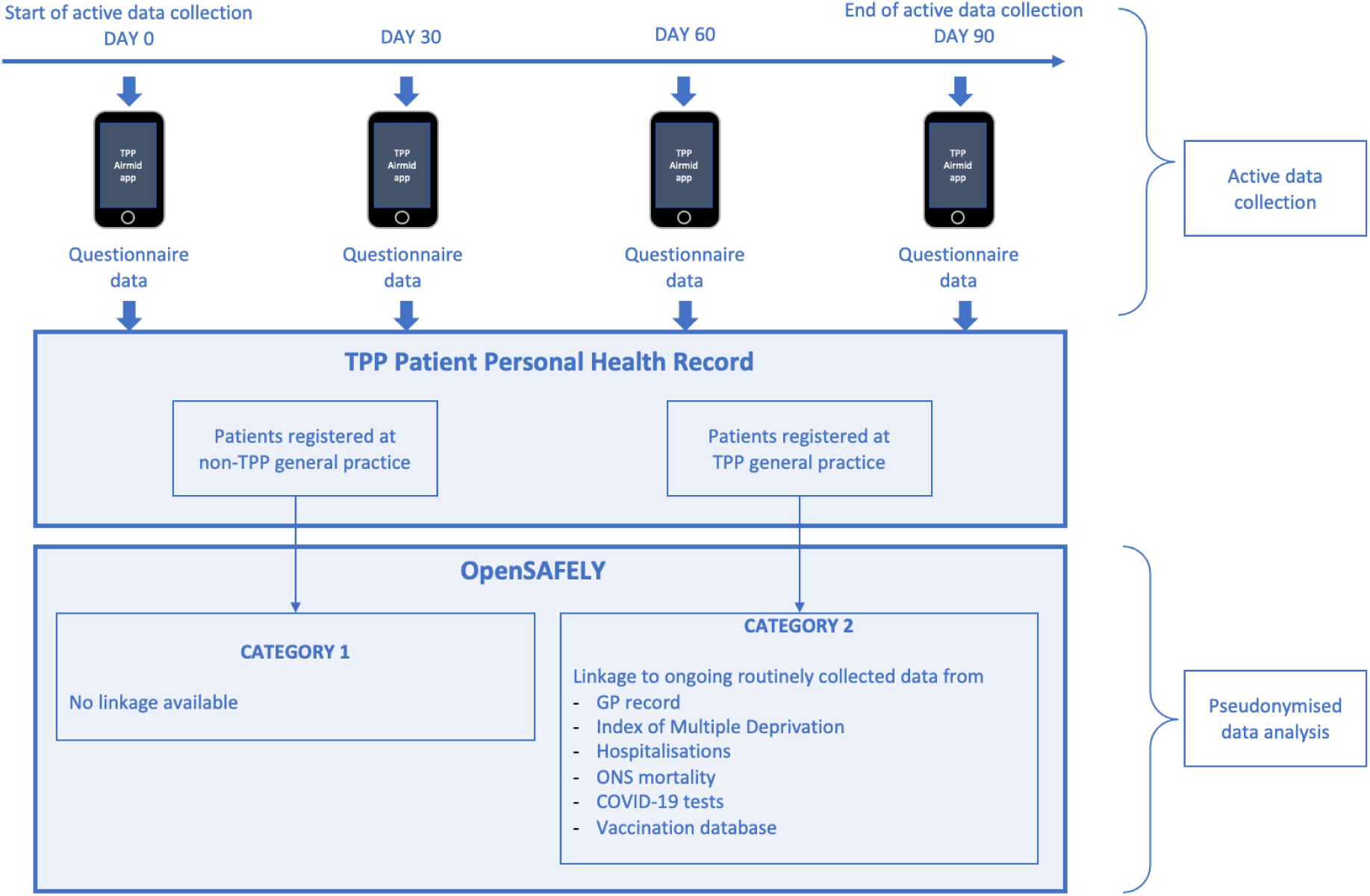
OpenPROMPT study design

OpenSAFELY provides a secure software interface allowing the analysis of pseudonymized primary care patient records from England, avoiding the need for large volumes of potentially disclosive pseudonymized patient data to be transferred off-site. This, in addition to other technical and organisational controls, minimises any risk of re-identification. Similarly pseudonymized datasets from other data providers are securely provided to the EHR vendor and linked to the primary care data. The dataset analysed within OpenSAFELY is based on 24 million people currently registered with GP surgeries using TPP SystmOne software. It includes pseudonymized data such as coded diagnoses, medications and physiological parameters. No free text data are included. Further details on our information governance can be found under “<u>Information governance</u>”.

To ensure protection of anonymity and in line with OpenSAFELY practice, we have censored any counts less than seven and have rounded all frequencies to the nearest 7 in this article. All code used for this report is freely available online at https://github.com/opensafely/openprompt-cohort-profile.

### OpenPROMPT cohort

The OpenPROMPT cohort comprises 7,574 individuals who have consented to data collection and responded to the baseline questionnaire, 6,337 of which proceeded to respond to our research questionnaire. We enrolled participants between November 2022 and July 2023 and each respondent was followed up for 90 days from baseline (data collection up to October 2023). We used the Airmid app to send notification reminders to respondents to complete the research questionnaire every 30 days. In total, 299,240 responses to our research questionnaire were collected over the study period (**Figure S1**).

Overall 4,599/7,574 (60.7%) of the baseline OpenPROMPT cohort identified as female and 7,203/7,574 (95.1%) reported their ethnicity as white. We collected information on the highest education level, relationship status, and household income at baseline. Amongst these categories approximately half of the cohort had attended college or university (4,004/7,574; 52.9%), were married or in a civil partnership (4,172/7,574; 55.1%), and had a household income between £26,000 and £63,999 (3,052/7,574; 40.2%) (**Figure 2B** & **Table 1**).

**Figure 2:**
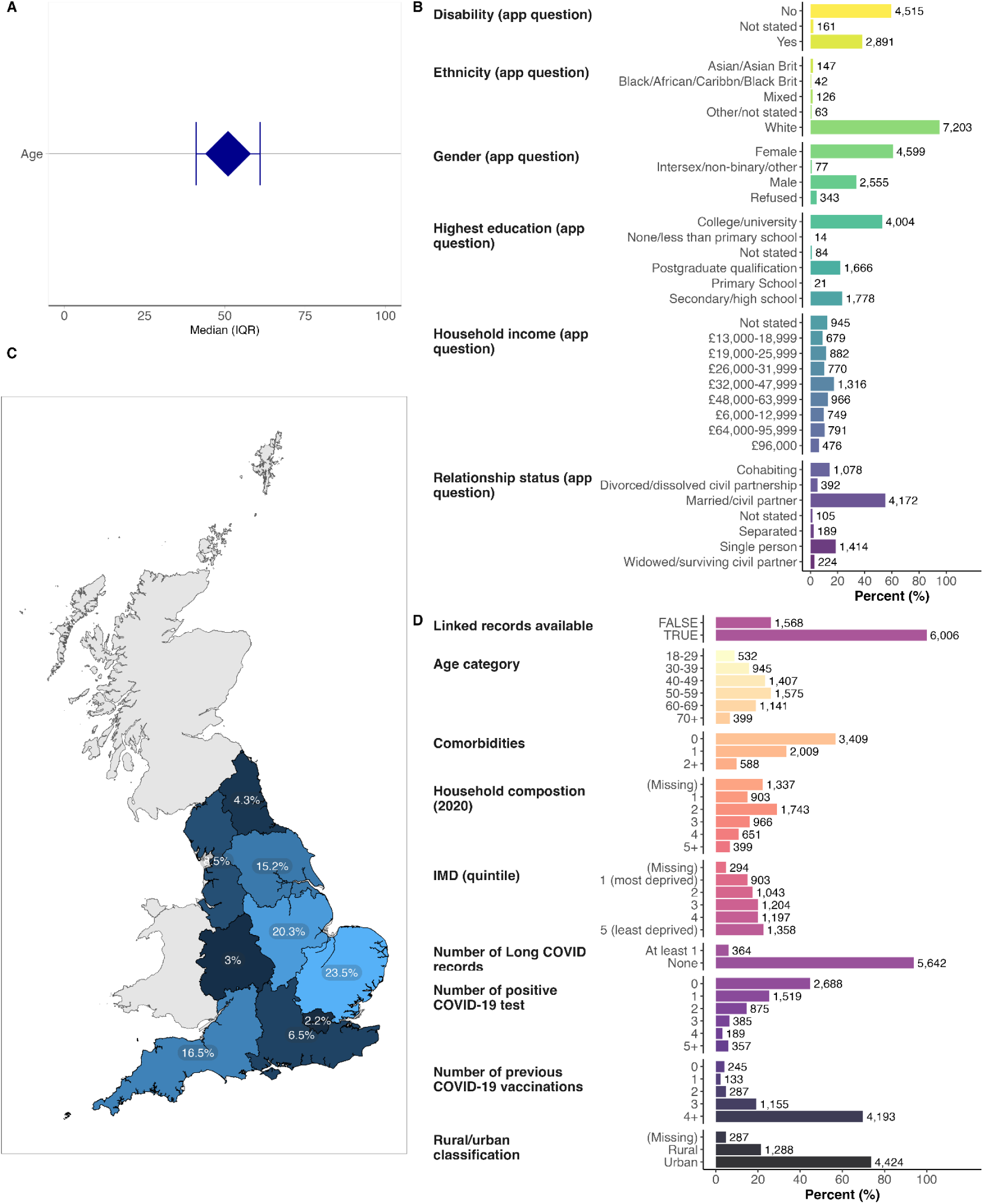
Baseline characteristics of the OpenPROMPT cohort. (A) median age and interquartile range for OpenPROMPT respondents with linked TPP data available (n=6,006). (B) Responses to the baseline OpenPROMPT questionnaire (n=7,574).(C) geographic distribution of respondents with linked TPP data available. It was possible for people to contribute to OpenPROMPT outside of England but linked data is available for English residents only. (D) further clinical and demographic data for the OpenPROMPT cohort with linked TPP data available (n=6,006)

We did not collect information on age in the app, but date of birth information is available in GP records linked with OpenSAFELY. Amongst the population with this linkage available (6,006; 79.3%) the median age was 51 (interquartile range: 41-61). We were able to analyse a wider range of demographic and clinical information in this subset with OpenSAFELY linkage available (**Figure 2C-D**). The largest representation of respondents was from the East of England (1,407; 23.5%) and East Midlands (1,217; 20.3%), but linked data was only available for respondents from England (**Figure 2C**). The majority of respondents were from urban areas (4,424/6,006; 73.7%) compared to rural areas 1,288 (21.4%) and information are available on household composition (collected in 2020) which showed that most OpenPROMPT respondents lived in household size of at least 2, although 903/6,006 (15%) respondents were living alone as of 2020.

We were also able to characterise the clinical status of participants with linked OpenSAFELY by investigating the clinical diagnoses in linked GP records. We counted the presence of previous morbidity codes for a range of common chronic conditions as in previous OpenSAFELY research including asthma, cancer, diabetes, cardiovascular disease and rheumatoid disease and other immunosuppressive conditions (full details in **Supplementary Methods**). We found that the majority did not have a comorbidity (3,409/6,006; 56.8%), but 2,009 (33.4%) had one recorded comorbidity and 588 (9.8%) had at least two comorbidities at baseline. Finally, we analysed linked records of the index of multiple deprivation and found that the most common quintile were the least deprived (1,358/6,006; 22.6%) and second least deprived (1,197; 19.9%) quintile, consistent with the reported household income information collected in the app (**Figure 2D**).

The advantage of linking data within OpenSAFELY is that we can directly compare the baseline characteristics of the OpenPROMPT cohort with the wider OpenSAFELY population of 24 million patients registered at TPP practices. We found that our cohort had greater representation of people aged between 40-70 but that there were fewer participants in younger and older age groups (**Figure 3A**). The OpenPROMPT cohort has a greater representation of individuals from less deprived areas, more female participants and more white participants than the overall OpenSAFELY population.

**Figure 3:**
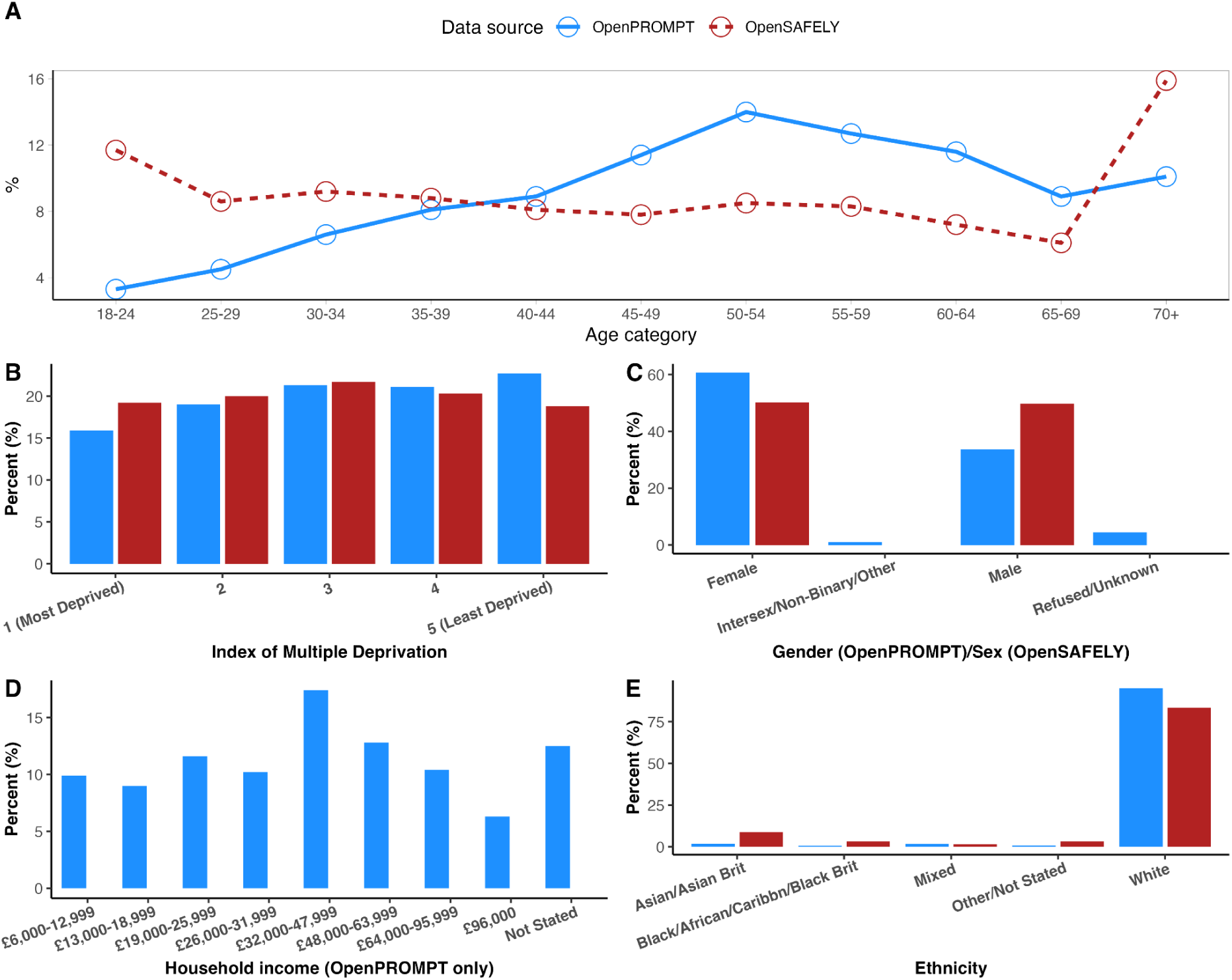
Comparison of OpenPROMPT cohort (blue) with the currently registered OpenSAFELY-TPP (red) population as of November 2022. OpenPROMPT data shown for those with a valid baseline response (n=7,574). In panel D, household income data is not collected by GPs so is not available in OpenSAFELY

### How often were they followed up and for how long?

The OpenPROMPT study was designed to collect information at baseline from respondents, and then every 30 days for a total of 90 days of follow up. Most OpenPROMPT participants were recruited in early 2023 and the final recruitment was in July 2023 (**Figure 4A**). Over the full 90 days of follow up, we collected a large number of responses to our baseline and research questionnaires, which were mostly from the first round of data collection (**Figure 4B**). We found that over half the cohort did not complete a follow up questionnaire at 30 days, and by 90 days we collected responses from 884 individuals (13.9% of original 6,337 respondents) (**Figure 4C**). It was possible for respondents to input data using the app at any point during follow up, so to retain consistency with our study protocol, we only included responses that were within a ±5 day window of the intended 30, 60, 90 days after the baseline questionnaire was completed, which means that some responses were discarded (**Figure 4D**).

**Figure 4:**
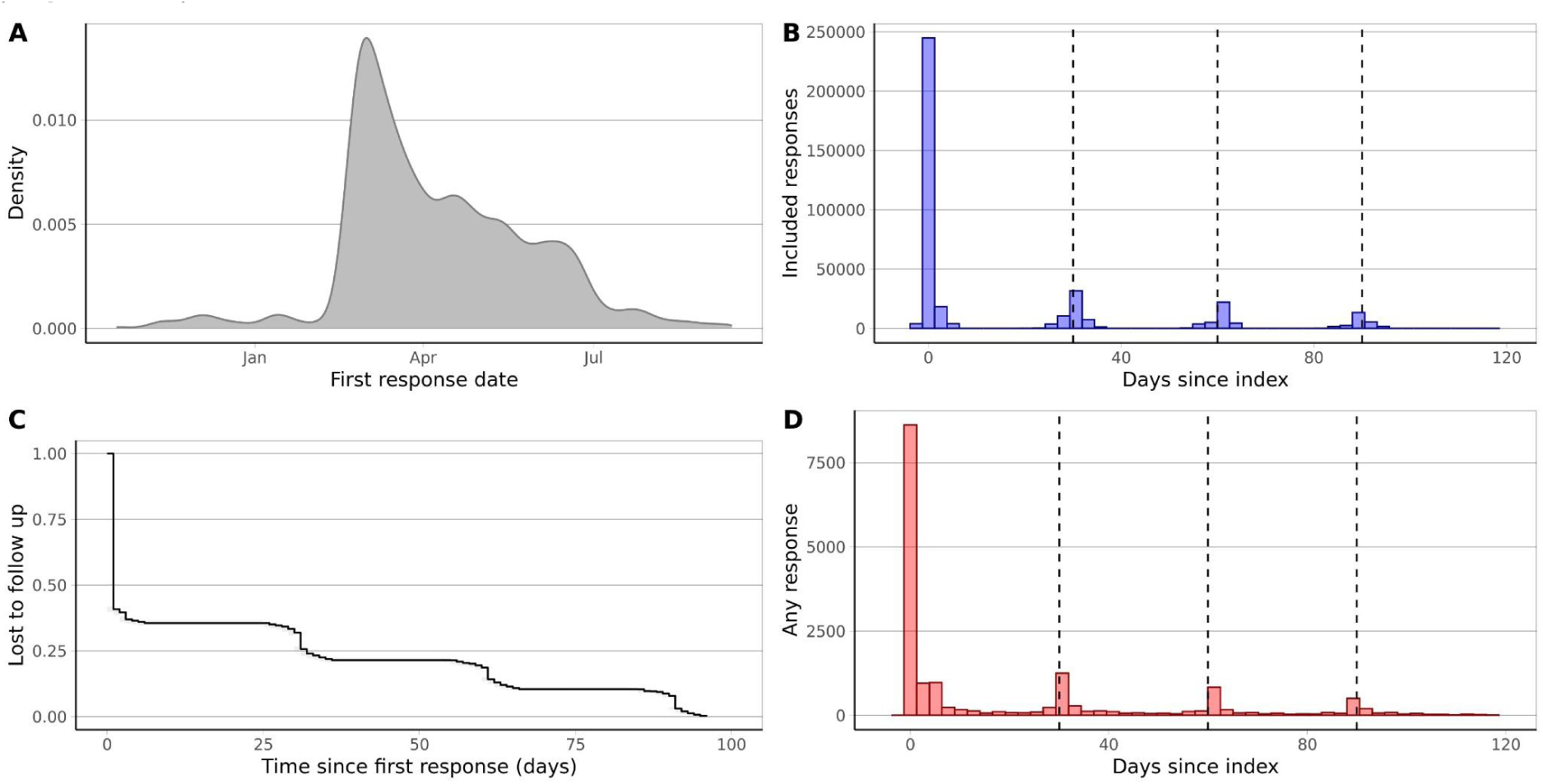
OpenPROMPT data collected over time. (A) density plot of the date of completion of the baseline questionnaire between November 2022 and July 2023. (B) Histogram of the number of valid responses to a question within OpenPROMPT over time. Only responses within a ±5 day window of the intended 30, 60, 90 days after the baseline questionnaire were included. (C) reverse Kaplan-Meier plot showing loss to follow up over time. The date of the last available data is shown along the x-axis. Time steps are shown once a total of 7 events occur to protect anonymity). (D) Histogram of the number of individuals that submitted a response over time. Responses not within the ±5 day window are excluded

### What has been measured?

We collected detailed data on the quality of life of the OpenPROMPT cohort including people living with and without long COVID. We recorded information from three standardised questionnaires: the Functional Assessment of Chronic Illness Therapy – Fatigue Scale (FACIT-Fatigue) (29), EQ-5D-5L (26), and the MRC breathlessness scale. We also asked questions about historic COVID-19 infections, the number and duration of symptoms, the vaccination status of respondents and whether or not they have recovered from their most recent COVID-19 infection.

Data collected with these questionnaires show a range of health related quality of life in the OpenPROMPT cohort. There was a notable proportion of people who reported severe fatigue using the FACIT questionnaire, especially for being “too tired to eat”, “feel weak” and “need help for usual activities” (**Figure 5A**). Similarly, many respondents had severe problems with anxiety/depression and pain/discomfort in the EQ-5D questionnaire (**Figure 5B**), and the most common MRC breathlessness grade was grade 2 (**Figure C**).

**Figure 5:**
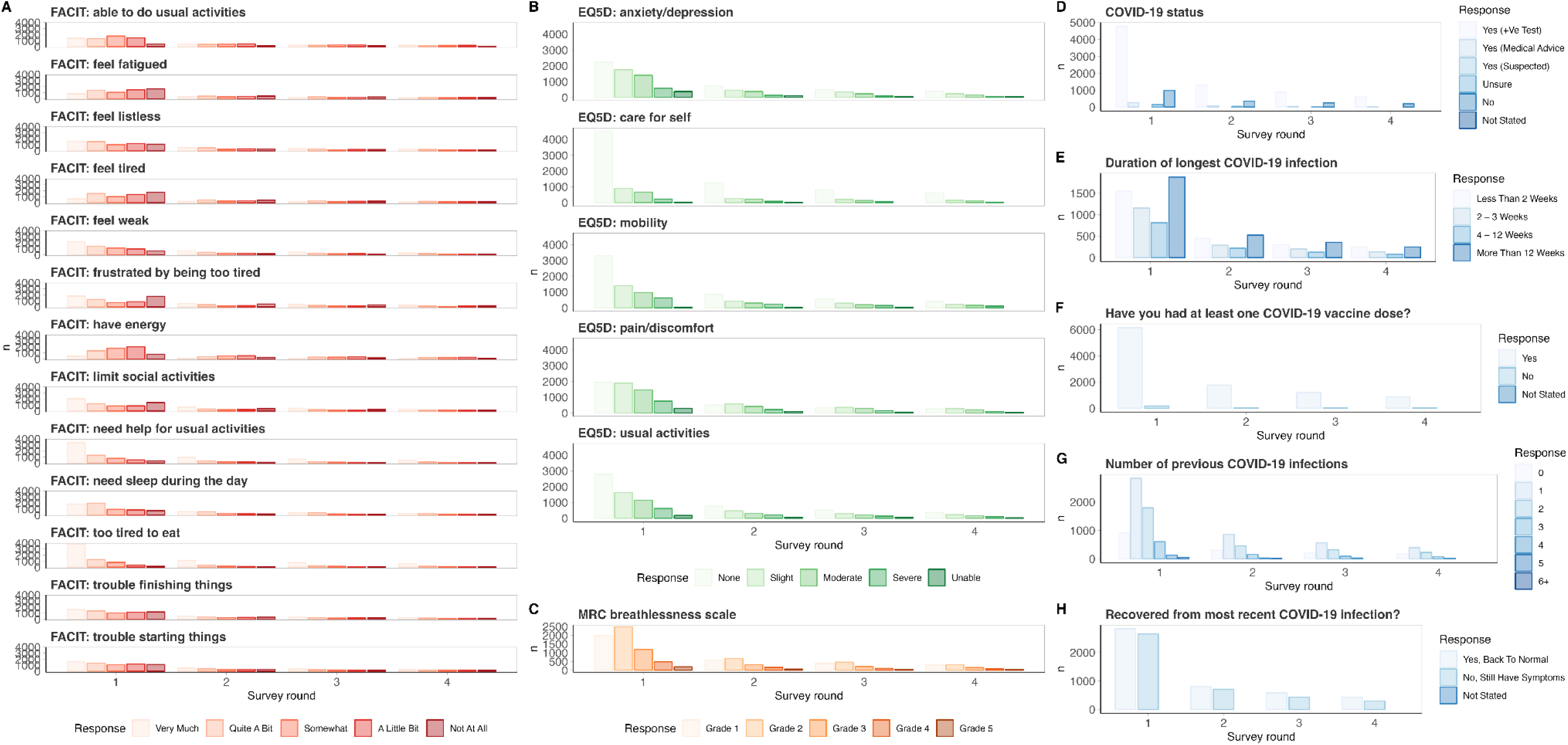
Questionnaire response in OpenPROPMT over 90 days from baseline. (A) Functional Assessment of Chronic Illness Therapy – Fatigue Scale (FACIT-Fatigue), (B) EQ-5D-5L, (C) MRC breathlessness scale, (D-H) responses to specific questions about COVID-19 infection history, duration of infection, SARS-COV-2 vaccinations and whether respondents had recovered from their most recent infection

We asked respondents about their experience with COVID-19, recovery and vaccinations. The vast majority of participants had a previous COVID-19 infection, mostly confirmed with a positive test (**Figure 5D**), and the most common duration of the longest COVID-19 infection was over 12 weeks. In the first survey round, only 1,547/5,390 (28.7%, the denominator is smaller because the question was optional) reported that the symptoms of their initial infection lasted less than 2 weeks (**Figure 5E**). Over 97% of the OpenPROMPT cohort had received a SARS-COV-2 vaccination dose at baseline (**Figure 5F**). In total, 6,337 people responded to the question “How many previous COVID-19 infections have you had?” during the first study round. 903 (14.3%) had not had a previous COVID-19 infection, almost half (2,842; 44.9%) had one previous infection and 784 had 3 or more infections previously (12.3%) (**Figure 5G**).

To assess the experience of long COVID in OpenPROMPT we collected relevant data from two questions. One asked about the duration of the longest infection with COVID-19 as described above. We also asked a more direct question, “have you recovered from your most recent COVID-19 infection?”. Across the study period we found that approximately half of respondents had not recovered from a COVID-19 infection. 2,653 of 5,488 responses at baseline reporting persistent symptoms (48.3%), and this proportion slightly declined over the 90 day study period to 46.8% at 30 days, 42.4% at 60 days and 40.8% at 90 days after baseline (**Figure 5H**).

We have demonstrated the advantage of collecting patient reported outcome measures linked to wider healthcare datasets above through direct comparison of the representativeness of our cohort. We can also compare the capture of healthcare outcomes in the two sources of data, since some questions in OpenPROMPT should also be collected in routine electronic health records (EHRs). We therefore compared the assessment of the same variables from different data sources in the same individuals with linked data available. We identified three questions regarding COVID-19 that have comparable data within OpenSAFELY. Firstly, the SARS-COV-2 vaccination status of respondents (“Yes” in **Figure 5F**) which we compared to any SARS-COV-2 vaccination records held within OpenSAFELY. Secondly, whether respondents had a previous COVID-19 infection (“Yes (+ve test)” in **Figure 5D**), which we compared to any positive result in the COVID-19 Second Generation Surveillance System (SGSS). Finally, we compared the proportion of people who were still experiencing symptoms in OpenPROMPT (“No, still have symptoms” in **Figure 5H**) with those with a GP record of long COVID, using a previously validated codelist for long COVID (16).

We found excellent agreement between OpenPROMPT and OpenSAFELY with respect to previous vaccination, demonstrating that the majority of vaccinations are well recorded in OpenSAFELY, and that information from this question is accurately captured in OpenPROMPT (**Figure 6A**). We also found very few examples (154; 3.2%) of people who had a record of previous COVID-19 infection in OpenSAFELY and did not report this in OpenPROMPT. However, a larger proportion of respondents did report a previous COVID-19 infection confirmed with a positive test in OpenPROMPT but had no record in OpenSAFELY (1,113; 22.9%) suggesting under-reporting of infections captured in SGSS, for example when lateral flow test results were not submitted online (**Figure 6B**). The worst agreement (Cohen’s Kappa = 0.15) was for recording of long COVID (**Figure 6C**). In OpenPROMPT 2,079 people reported persistent symptoms at baseline, and had linked data available. However, only 315 (15.1%) also had a GP record for long COVID (7.3% of the total linked data sample).

**Figure 6:**
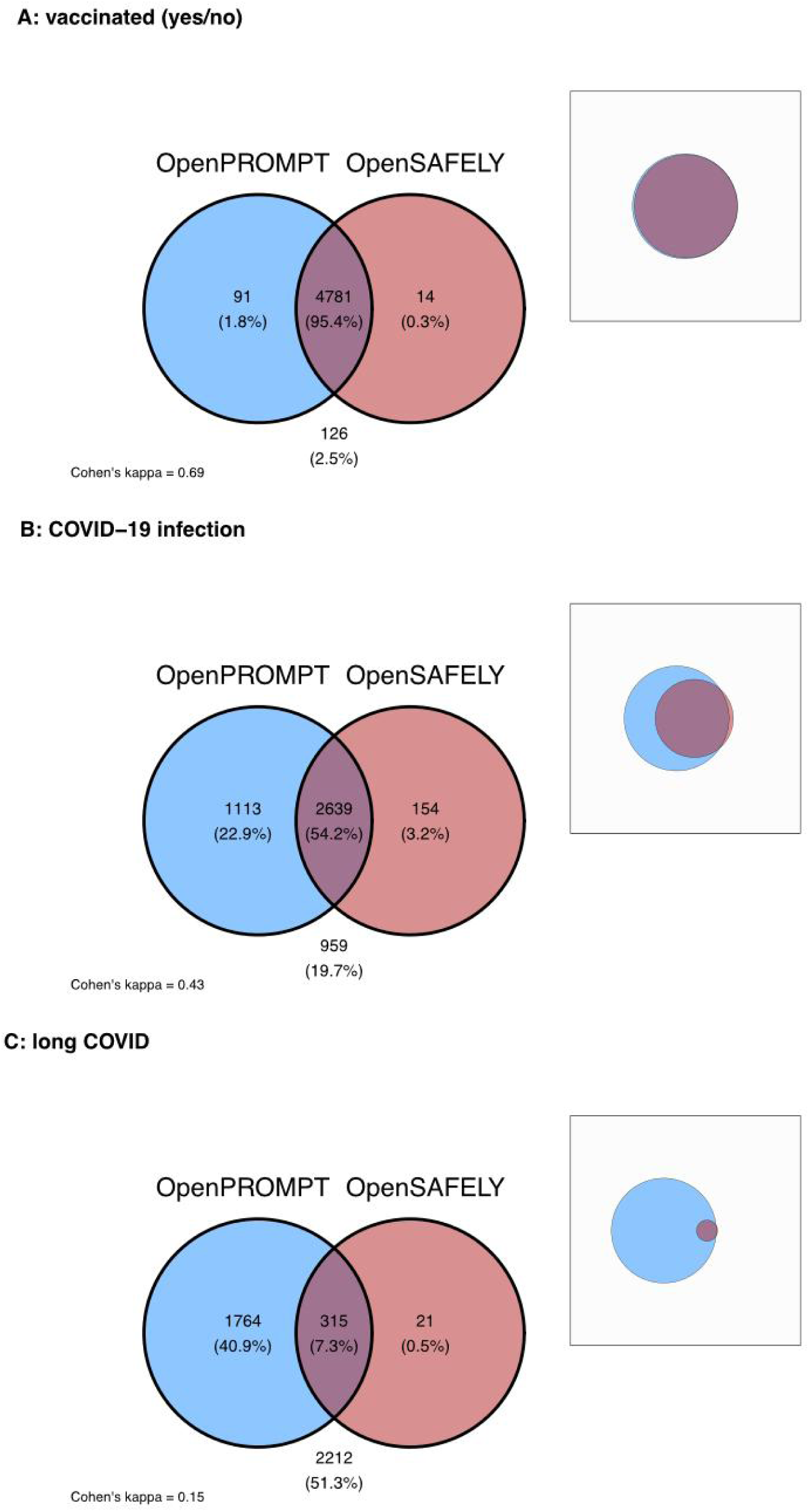
Venn diagrams demonstrating agreement between patient reported (OpenPROMPT, blue) and routinely collected (OpenSAFELY, red) data for the same clinical information in individuals with linked data. Frequency and percentage of total are shown for each variable, the inset shows these proportions on a relative scale. (A) previous vaccination, (B) previous Covid-19 infection, (C) unresolved persistent symptoms following Covid-19 infection

## How can the data be accessed?

The OpenPROMPT questionnaire data provided by participants will be made available to allow other researchers to benefit from this work in the future, and for a range of different studies and purposes. Data access will be free, please contact. If accessing through OpenSAFELY then access will be subject to completion of data security requirements specified by the OpenSAFELY platform at the time of application. All other study documentation will be stored for a minimum of 5 years after the follow-up period.

## What are the main strengths and weaknesses?

The main strength of OpenPROMPT is that we have collected data on HRQoL using validated instruments, especially the EQ-5D, in people with and without long COVID, and linked these data to a large EHR resource. This facilitates a much wider range of research including information on healthcare history, prescriptions, COVID-19 vaccinations and other healthcare data, without participants having to answer excessive questionnaires, which is especially pertinent in long COVID research.

Linkage between OpenPROMPT and OpenSAFELY is also valuable from an epidemiological perspective as it facilitates direct data comparisons between two separate sources. This means that we can investigate which routinely collected variables are well recorded, for example Covid-19 vaccinations, and which ones are not, such as recovery from long COVID symptoms.

Finally, We embedded patient and public involvement (PPI) throughout the development and delivery of OpenPROMPT through three mechanisms. Firstly, OpenPROMPT recruited a three person steering group, including members with long COVID which met with the research team online every 6 months. Secondly, we held two separate PPI events in January and September 2023 to obtain feedback on the study and respond in real time. Finally, engagement with research using electronic health records was also being undertaken as part of OpenSAFELY. This includes formation of an Oversight Board (which reviewed OpenPROMPT) (https://www.opensafely.org/governance/), publishing blogs explaining OpenSAFELY (https://www.opensafely.org/blog/), presenting results of studies at public and professional events, soliciting feedback on studies and conducting focus groups.

There are limitations in representation of our OpenPROMPT sample with regards to certain demographic variables. OpenPROMPT includes an over-representation of women, those of white ethnicity and less deprived areas as measured by IMD compared to the average in the wider OpenSAFELY population. This, in part, reflects the overrepresentation of people with long COVID in OpenPROMPT, which has been reported higher in these same groups, however this does need to be considered when analysing OpenPROMPT data. In particular, our study was susceptible to digital exclusion as users required a smartphone and the ability to use it confidently in order to join the study. This digital exclusion will disproportionately impact certain groups in the population from which the sample has been drawn. The generalisability of findings from OpenPROMPT data to the wider UK population are therefore uncertain, for example we are unable to estimate the prevalence of long COVID in the UK from these data.

We also experienced a relatively high loss to follow up during the study. We aimed to reduce the burden of completing these questionnaires, especially since our study focussed on people living with a chronic condition which impacts fatigue. We made the questionnaires available in an app and it was possible to save progress and return at a later time. However, the high rate of loss to follow up does suggest that there was a selection pressure from our data collection method and this should be recognised in analysis of the longitudinal OpenPROMPT data. There are likely to be systematic differences between those that do and do not complete follow up and so future studies should include plans to improve retention over the planned study period, since the convenience of a smartphone app was insufficient in our experience.

## Administrative

### Acknowledgments

We are very grateful for all the support received from the TPP Technical Operations team throughout this work, and for generous assistance from the information governance and database teams at NHS England and the NHS England Transformation Directorate.

### Information governance

LSHTM is the data controller of OpenPROMPT data. NHS England is the data controller of the NHS England OpenSAFELY COVID-19 Service; TPP is the data processor; all study authors using OpenSAFELY have the approval of NHS England (30). This implementation of OpenSAFELY is hosted within the TPP environment which is accredited to the ISO 27001 information security standard and is NHS IG Toolkit compliant (31); Patient data has been pseudonymised for analysis and linkage using industry standard cryptographic hashing techniques; all pseudonymised datasets transmitted for linkage onto OpenSAFELY are encrypted; access to the NHS England OpenSAFELY COVID-19 service is via a virtual private network (VPN) connection; the researchers hold contracts with NHS England and only access the platform to initiate database queries and statistical models; all database activity is logged; only aggregate statistical outputs leave the platform environment following best practice for anonymisation of results such as statistical disclosure control for low cell counts (32).

The service adheres to the obligations of the UK General Data Protection Regulation (UK GDPR) and the Data Protection Act 2018. The service previously operated under notices initially issued in February 2020 by the the Secretary of State under Regulation 3(4) of the Health Service (Control of Patient Information) Regulations 2002 (COPI Regulations), which required organisations to process confidential patient information for COVID-19 purposes; this set aside the requirement for patient consent (33). As of 1 July 2023, the Secretary of State has requested that NHS England continue to operate the Service under the COVID-19 Directions 2020 (34). In some cases of data sharing, the common law duty of confidence is met using, for example, patient consent or support from the Health Research Authority Confidentiality Advisory Group (35).

Taken together, these provide the legal bases to link patient datasets using the service. GP practices, which provide access to the primary care data, are required to share relevant health information to support the public health response to the pandemic, and have been informed of how the service operates.

### Ethics

This research is part of the OpenPROMPT study “Quality-of-life in patients with long COVID: harnessing the scale of big data to quantify the health and economic costs” which has ethical approval from HRA and Health and Care Research Wales (HCRW) (IRAS project ID 304354).

### Ethical review

The Study Coordination Centre has obtained approval from the LSHTM Research Ethics Committee (ref 28030), as well as a favourable opinion from the South Central—Berkshire B Research Ethics Committee (ref 22/SC/0198).

### Dissemination plan

All publications and presentations relating to the study will be authorised by the study management group and also follow the OpenSAFELY publication policy. All publications resulting from the OpenPROMPT study will be open access.

### Data sharing

The questionnaire data provided by participants will be made available to allow other researchers to benefit from this work in the future, and for a range of different studies and purposes. Opting-in to the study required an affirmative response to agree to use of data in this way. After completion of the study, a pseudonymised copy of the data will be held according to NHS England retention policy. Data access will be governed by LSHTM and will require researchers to complete a data access form. All other study documentation will be stored for a minimum of 5 years after the follow-up period.

## Appendix

### Supplementary Methods

Comorbidities were assessed at study entry. Relevant comorbidities were defined based on previous research of risk factors for Long COVID and COVID-19 in OpenSAFELY (1–3). A previous code for one or more of: diabetes; cancer; haematological cancer; asthma; chronic respiratory disease; chronic cardiac disease; chronic liver disease; stroke or dementia; other neurological condition; organ transplant; dysplasia; rheumatoid arthritis, systemic lupus erythematosus or psoriasis; or other immunosuppressive conditions. Those with no relevant code for a condition will be assumed not to have that condition. Number of conditions were categorised into “0”, “1”, and “2 or more”. All codelists used to define these comorbidities are listed on GitHub (https://github.com/opensafely/openprompt-cohort-profile/tree/main/codelists), and are publicly available on https://www.opencodelists.org/.

### Supplementary Figures

**Figure S1:**
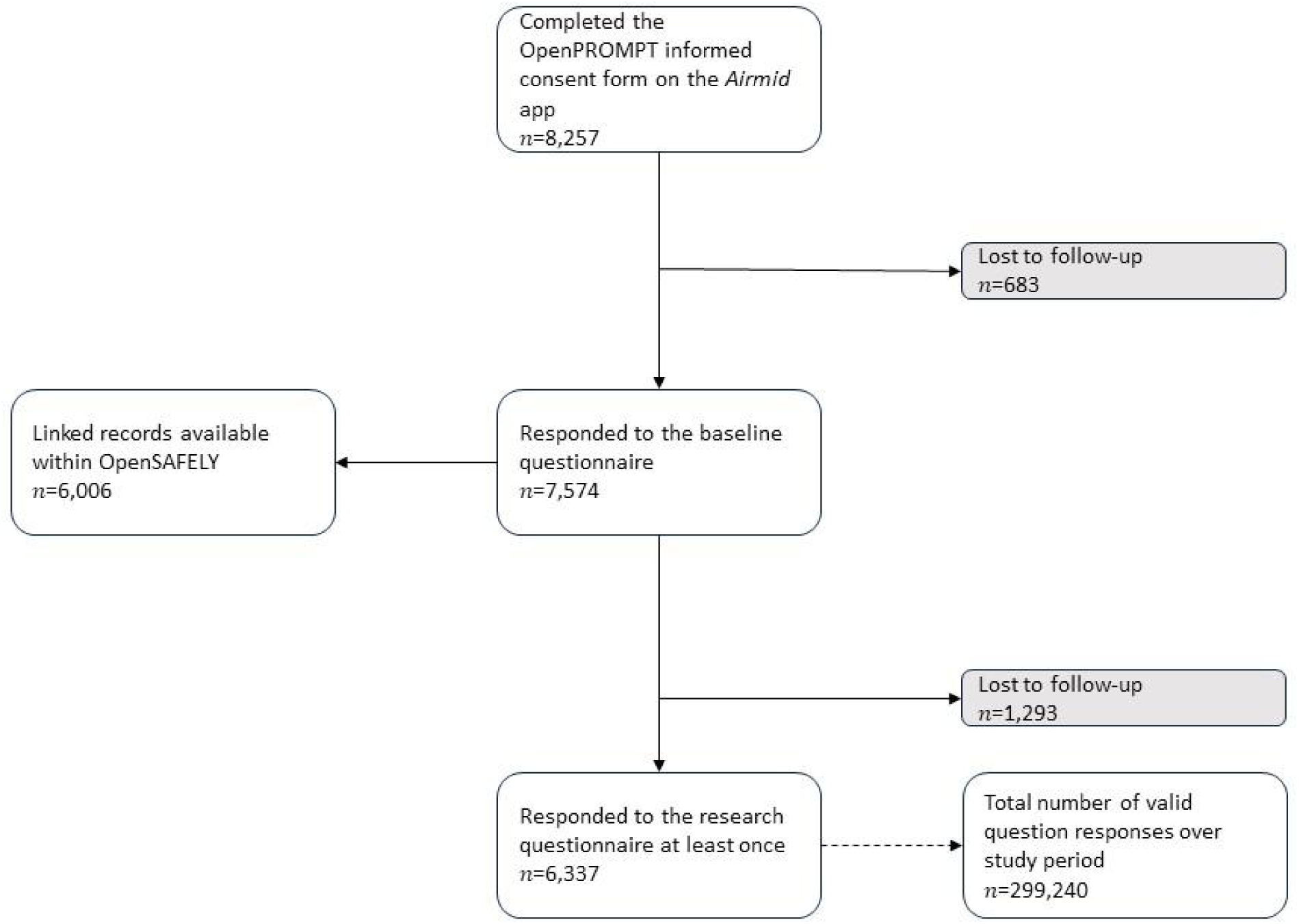
Cohort enrolment using an app-based data collection method for OpenPROMPT

### Supplementary Tables

**Table S1:**
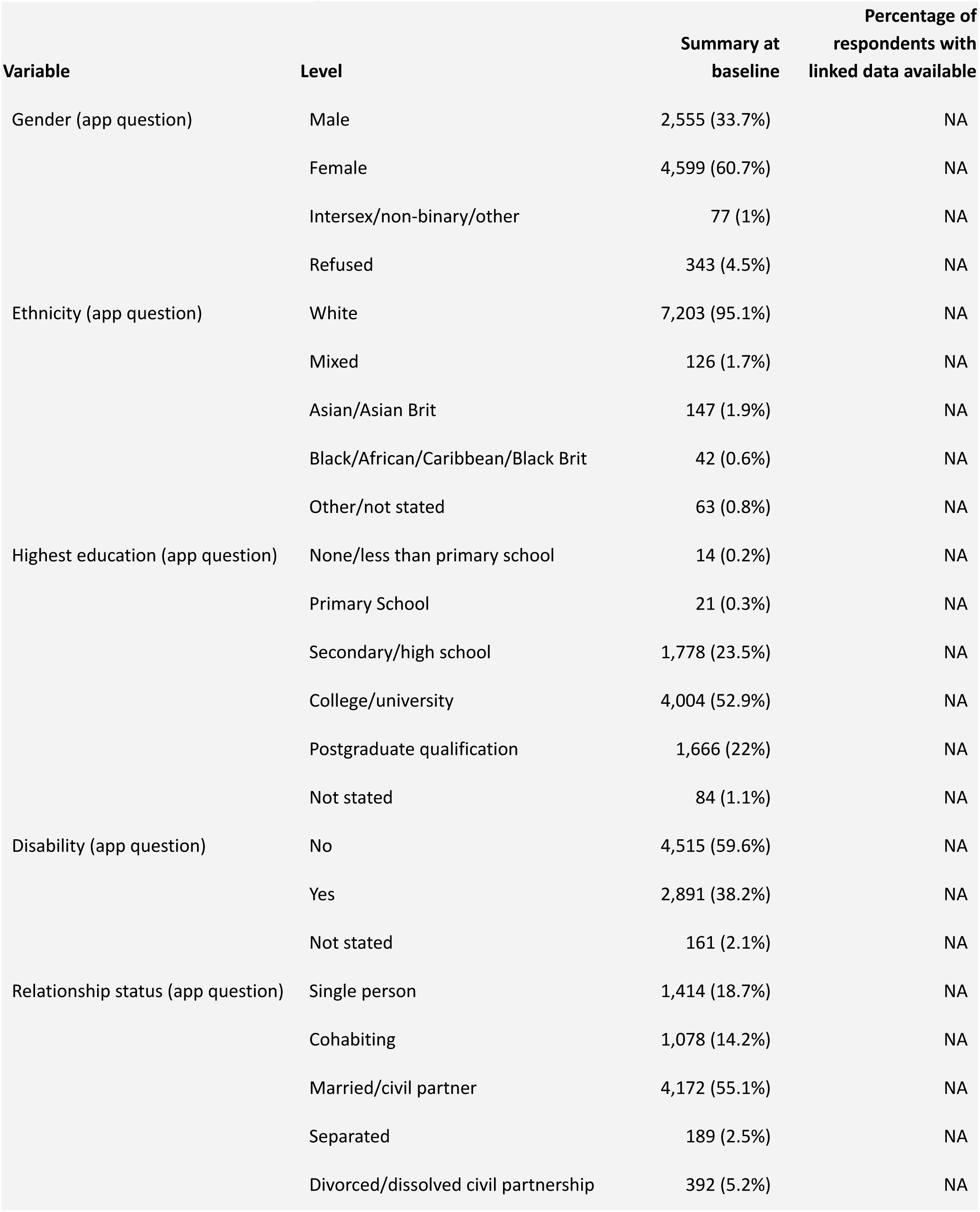
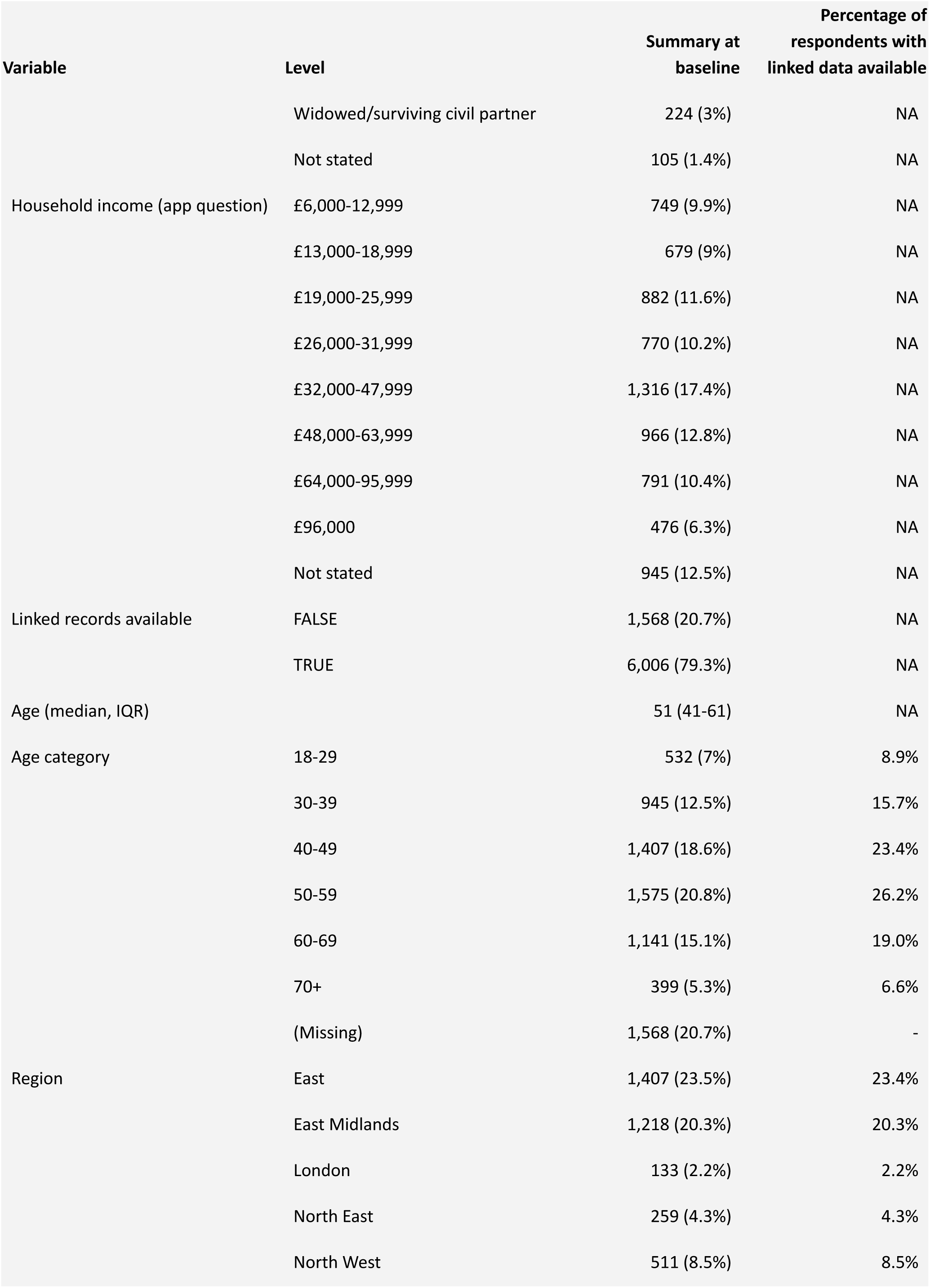
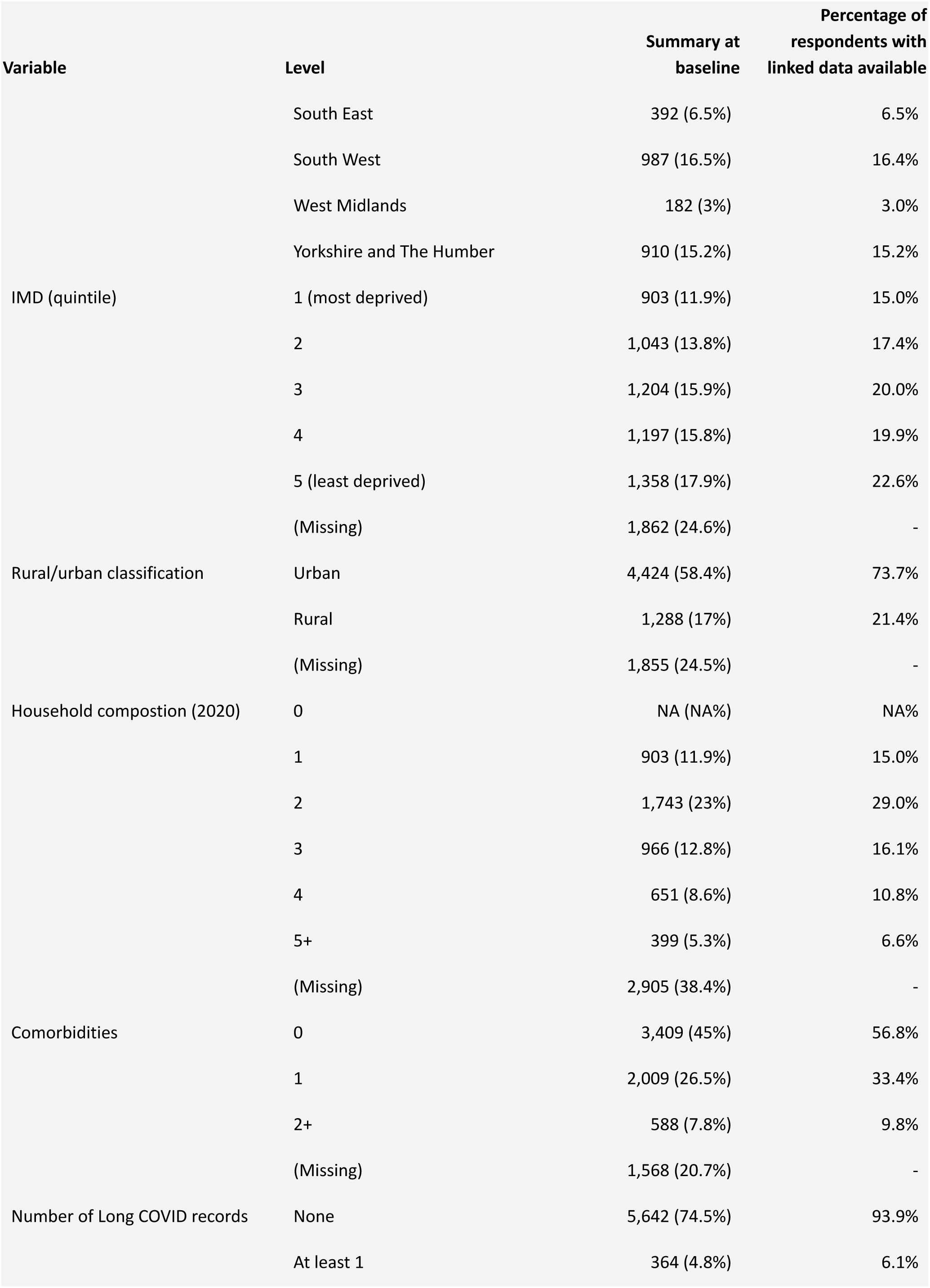
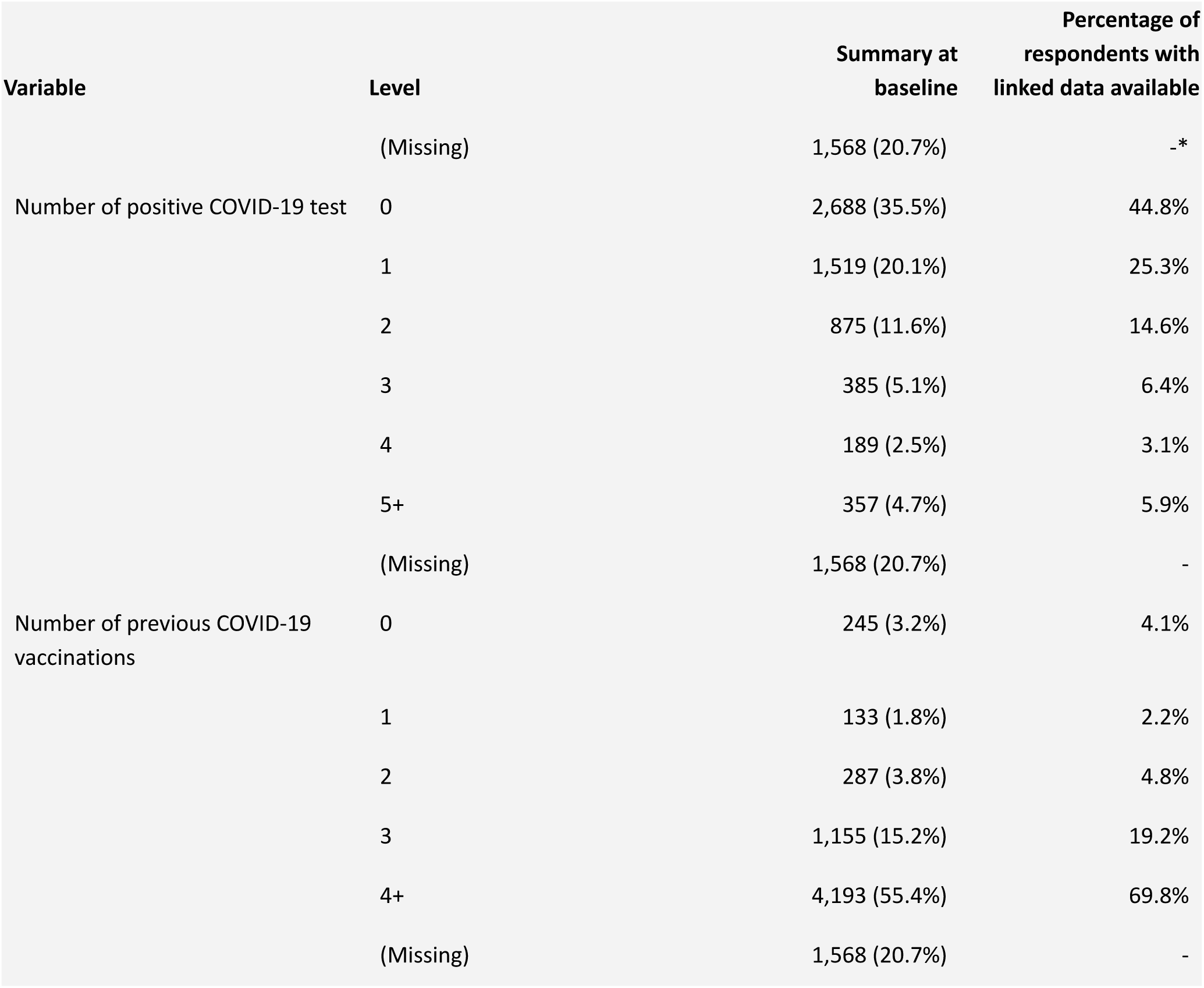
characteristics of OpenPROMPT participants at cohort entry and responses to the baseline questionnaire

**Table S2:**
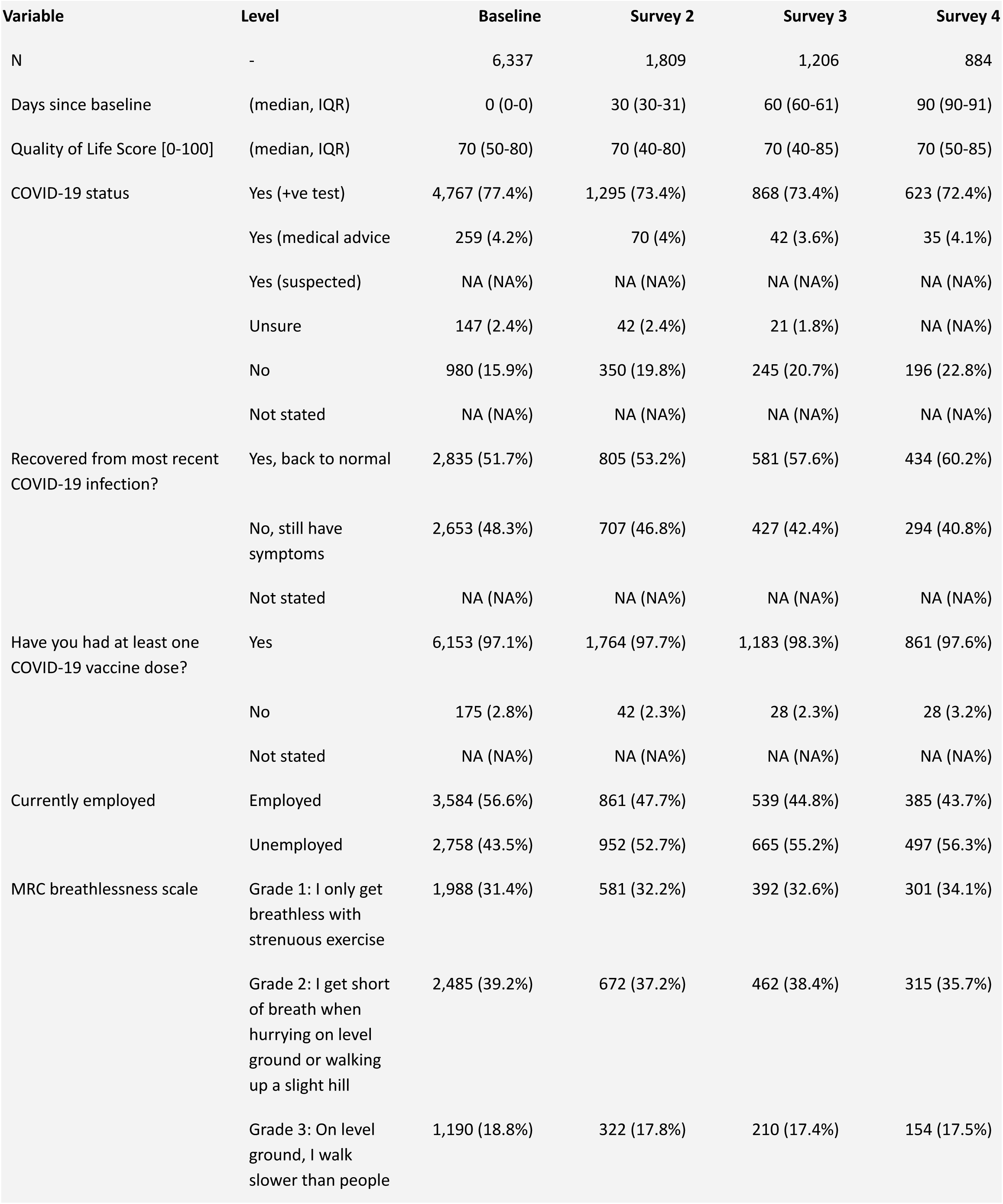
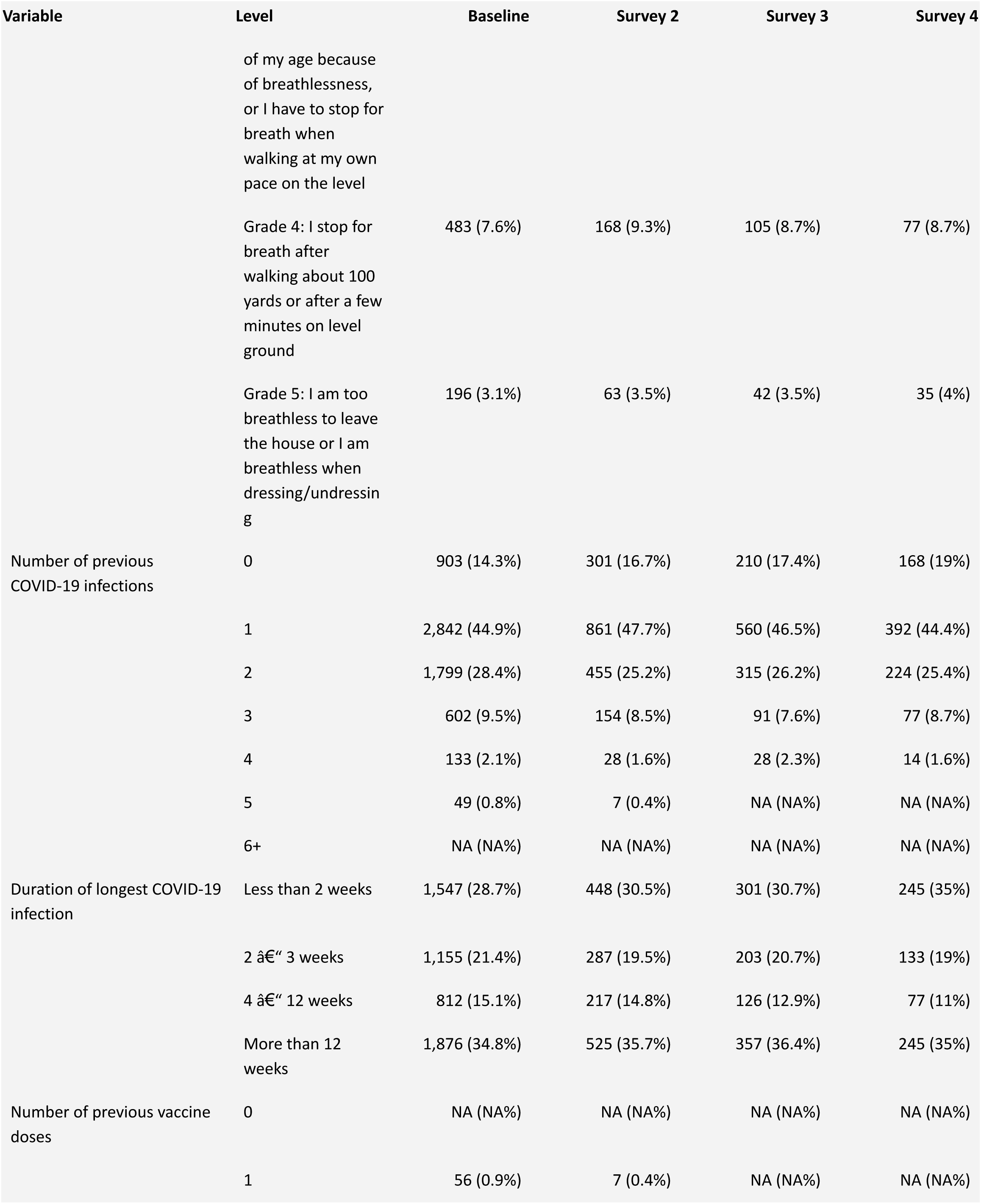
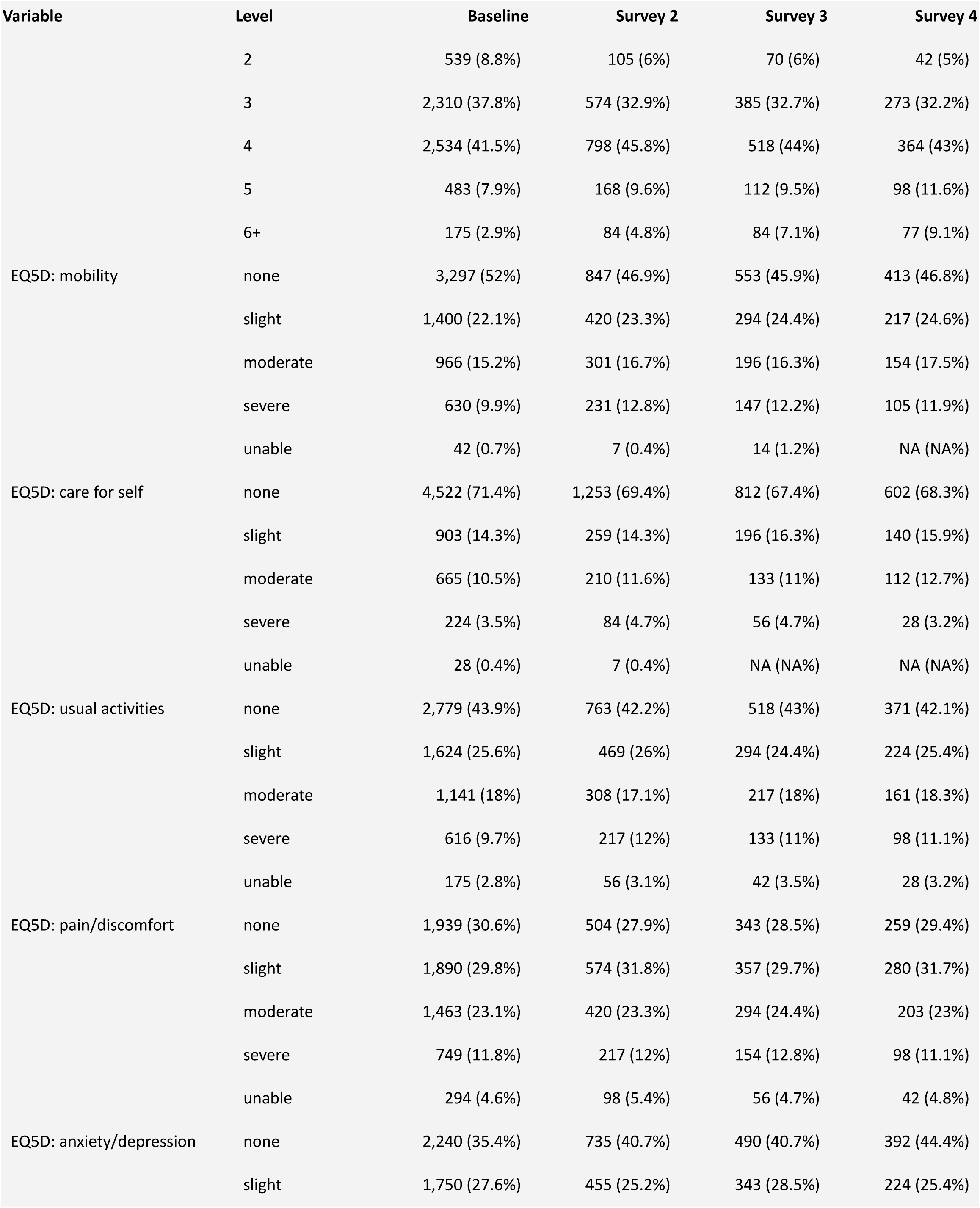
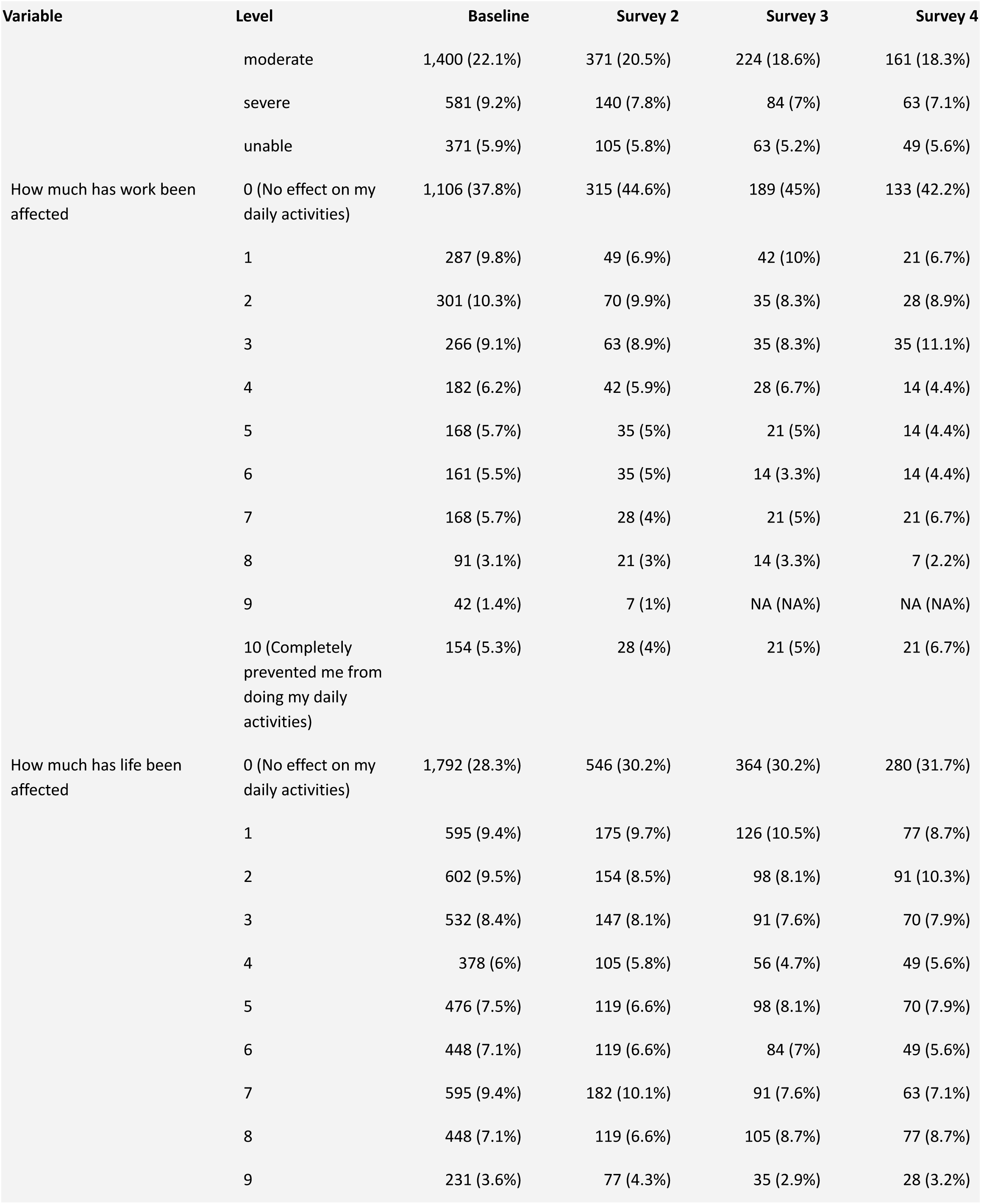
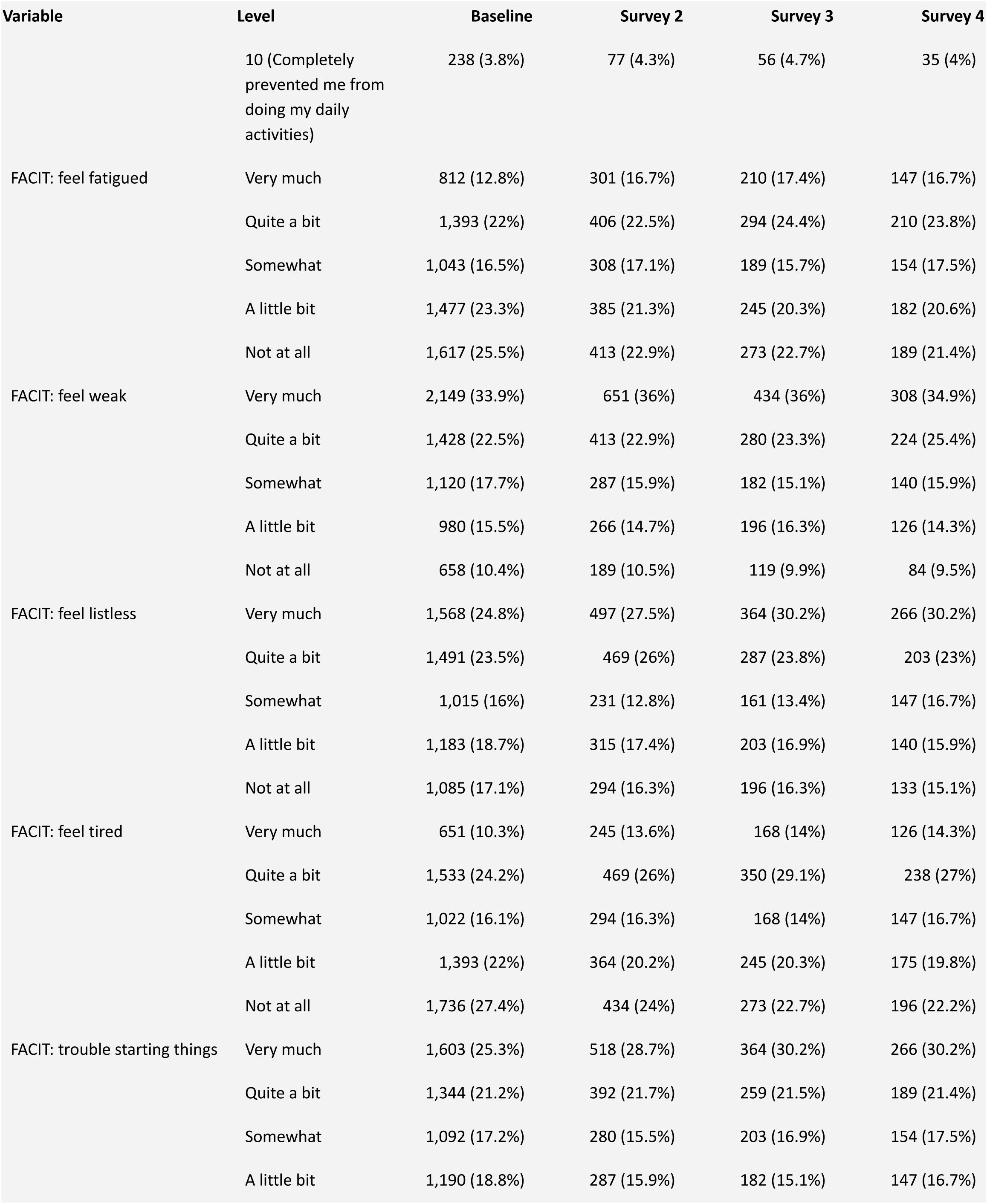
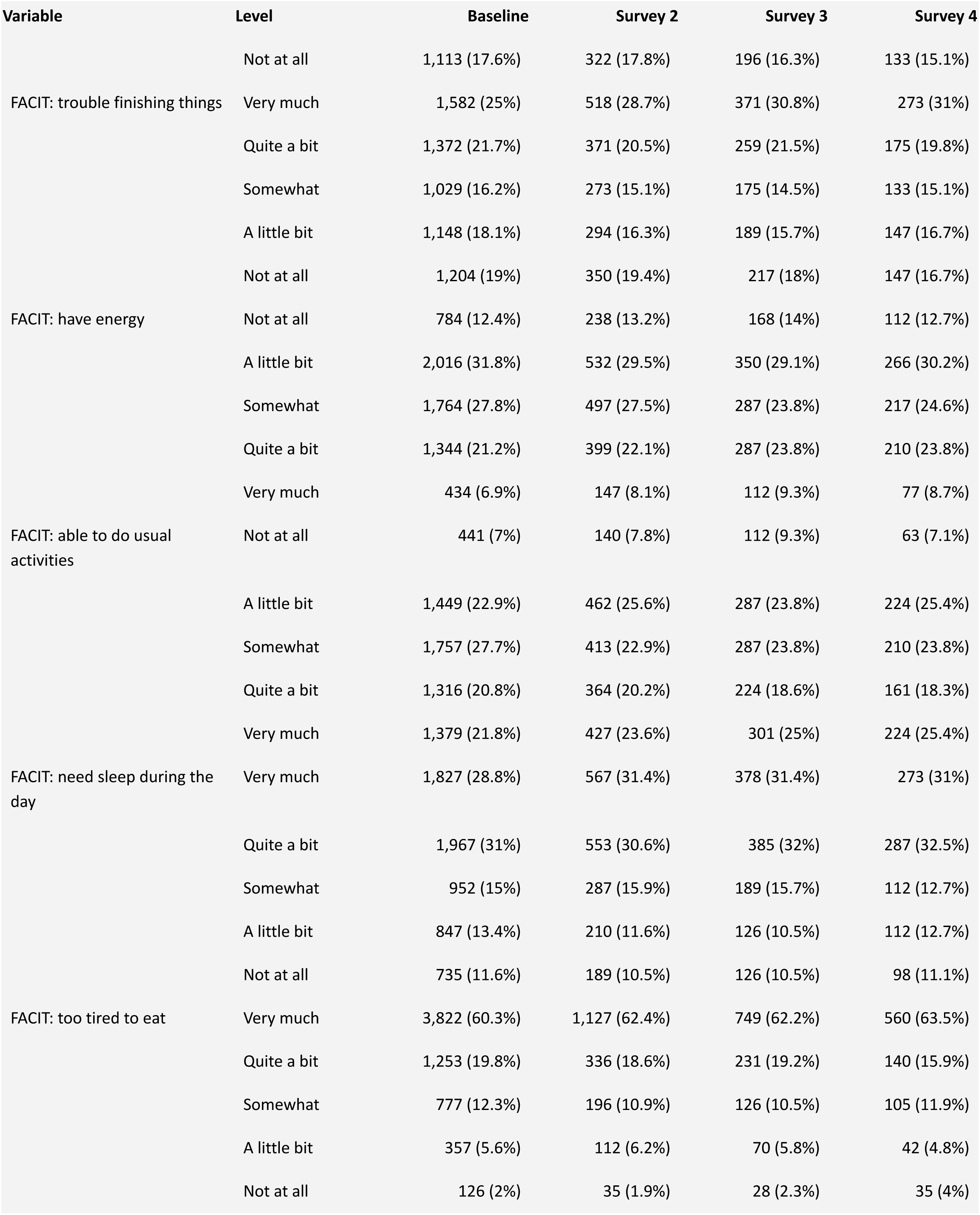
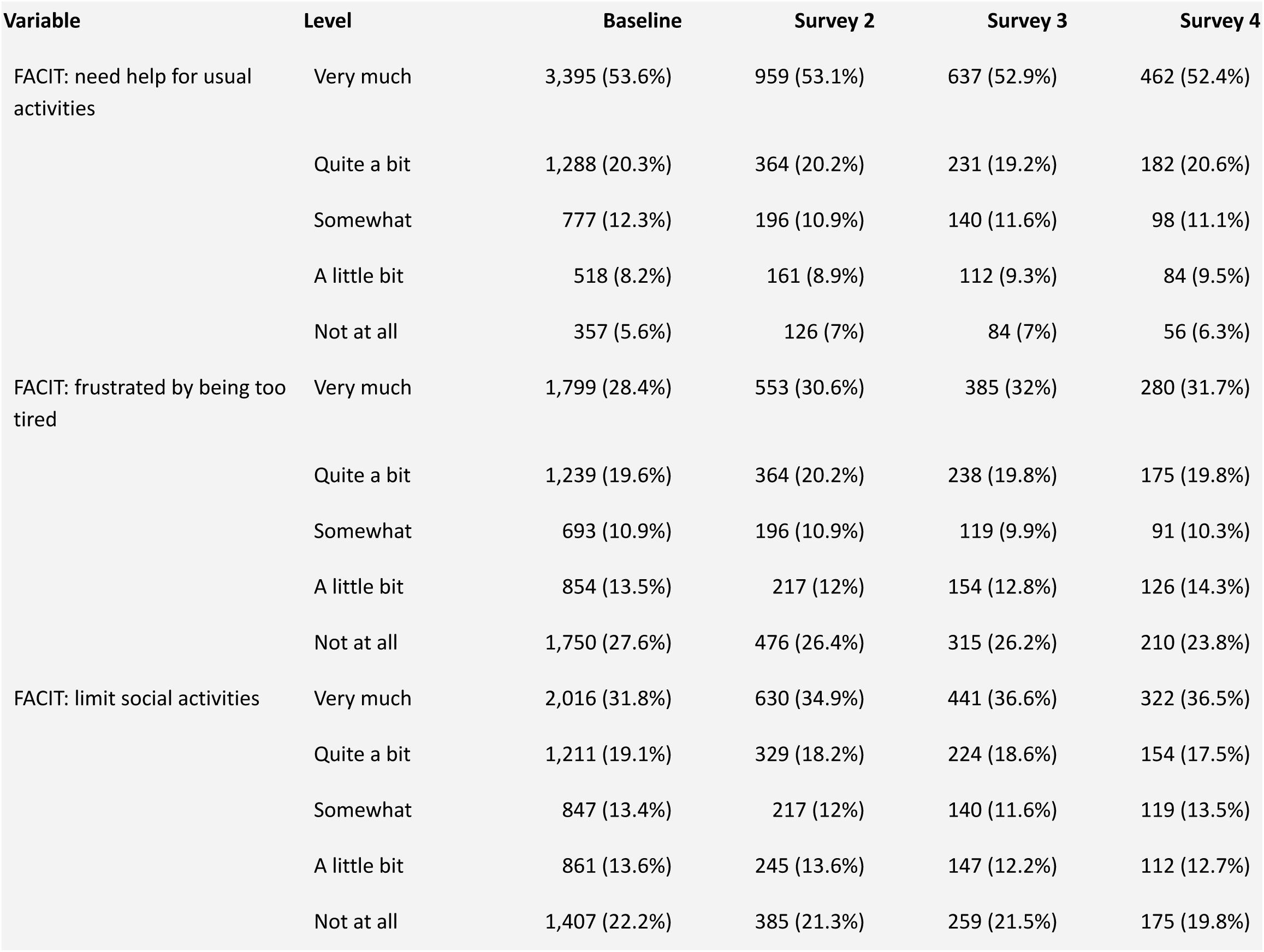
Responses to the OpenPROMPT research questionnaire over time since study enrolment (baseline, +30 days, +60 days, +90 days)

